# Day-to-Day Activity Avoidance in the Context of Urinary and Fecal Incontinence: Preliminary Results from Young Adults with Spina Bifida in an Ecological Momentary Assessment Study

**DOI:** 10.1101/2025.05.20.25327607

**Authors:** Devon J. Hensel, Audrey I. Young, Konrad M. Szymanski

## Abstract

**Background:** Studies suggest that young adults with spina bifida (YASB) often avoid important daily activities (ADL) due to urinary (UI) and fecal (FI) incontinence, but no research has prospectively examined how often ADL avoidance occurs, or the context in which it occurs. We used ecological momentary assessment (EMA) over 30 days to describe the frequency of avoidance, the activities YASB most commonly avoid, and the preliminary association of daily avoidance with affect, incontinence anxiety and health-related quality of life (HRQoL).

**Method:** We analyzed a subsample of YASB participants (18 – 27 years; N=23 of total 88 participants) who completed a larger 30-day prospective EMA study examining the daily prevalence and context of UI and FI in adults with SB. Participants completed an end-of-day EMA tracking daily ADL avoidance on days when they were worried about possible UI or FI and on days when the had actual UI or FI. Additional day-level measures were affect and incontinence anxiety. HRQoL was reported in beginning- and end-of-study surveys.

**Results:** YASB contributed 643 total EMAs; 57.5% had any incontinence. ADL avoidance was most frequent with actual FI (20.5%) and least frequent with worry about FI (3.5%, p=0.02) (UI worry: 8.3; actual UI: 8.9%, p=0.546). The most common basic ADL avoided were eating, drinking and spending time with family/friends. Negative mood and incontinence anxiety were significantly higher on days with all types of avoidance. Higher baseline HRQoL was associated with fewer in-study ADLs avoided, which were in turn associated with higher end-of-study HRQoL.

**Discussion:** YASB avoid key activities of daily living (e.g., eating, drinking, seeing family/friends) both when they worry about possible incontinence that may but does not occur, and when they experience actual incontinence. In other words, a dry day is not a day free of the negative effects of incontinence. ADL avoidance patterns vary between UI and FI. Tailored interventions addressing actual incontinence and worry about possible incontinence incorporating daily mood or incontinence anxiety can potentially minimize ADL avoidance and its impact on long-term HRQoL.

**Discussion and Implications:** YASB avoid key activities of daily living (e.g., eating, drinking, seeing family/friends) both when they worry about possible incontinence and when they experience actual incontinence. Tailored intervention may consider addressing daily mood or incontinence anxiety to minimize any ADL avoidance in the short term as a means to improve HRQoL in the long term.

## Background

Urinary (UI) and fecal (FI) incontinence are common secondary health conditions among young adults with spina bifida (YASB). Large scale studies report past month UI prevalence between 50% and 75% and past month FI prevalence between 41% and 55%.^1^ Many YASB report both UI and FI in the past month.^2^ Experiencing *actual* UI or FI and/or worrying about *possible* UI or FI can generally interfere with YASB’s participation in different activities of daily living (ADL).^3-5^ ADL are routine but essential tasks that individuals must be able to perform without assistance in order to be able to live independently.^6^ People born with SB now have a longer lifepan,^7^ meaning that YASB will live with UI and FI for several decades. Because of this increased lifespan, SB care guidelines now recommend that YASB to self-manage as many ADL as possible, as early as possible.^8,9^ Maximizing ADL participation as YASB live amidst actual incontinence or worry about possible incontinence offers an important means for them to integrate themselves – even for YASB who are partially or completely dependent on caregivers – more fully into society at large.^9^ In addition, greater ADL independence could facilitate the transition to adult care for some YASB^10^ and is likely to bolster health related quality of life (HRQoL) for all YASB.^11^ As with many other chronic illnesses, both increased independence and increased HRQOL are central therapeutic goals for YASB.

Providing support to YASB to reduce any incontinence-related ADL avoidance relies upon capturing what “clinically significant” avoidance looks like in any given YASB’s daily life. Clinical experience and our own research data support the idea that UI and FI vary not only from person to person, but also within the same person over different days.^2,12-15^ Yet, basic questions about daily experiences needed to support clinical decision making – for example, *how often* does ADL avoidance occur in daily life, and *which activities* are most commonly avoided? Do these factors *differ between UI and FI*, whether an individual *experiences actual incontinence or only worries about incontinence that may but does not occur*, and do they *differ depending on the context of the day*? – remain unaddressed in the extant literature. Moreover, our past cross-sectional work suggests that objective aspects of UI and FI are independently associated with lower HRQoL,^1,16,17^ but it is unclear to what extent higher HRQoL may reduce daily ADL avoidance, and to what extent higher rates of daily ADL avoidance may reduce later HRQoL. Capture of these data is therefore an essential part of personalized continence care, as ADLA therapies can be directly tailored from YASB’s own reports of their “real life” experiences.^12^

In the current paper, we use ecological momentary assessment (EMA) to gather these necessary data. Traditional measurement relies on YASB to retrospectively self-report incontinence experiences over a long interval (e.g., “in the past month” or “since the last clinic visit”).^2^ Data elicited from these approaches can introduce measurement bias, providing a less-than-accurate observation of ADLA in “everyday life.”^18-21^ In contrast, EMA is a prospective – or forward looking – data collection approach that reduces this measurement bias through brief-but-repeated “in the moment” reports of YASB’s daily experiences (e.g., ADL avoidance due to worry about or actual UI or FI) and the context (e.g., mood or feelings about incontinence) in which they occur.^22^ Over time, these repeated snapshots coalesce to provide a more accurate portrait of ADLA occurrence *in real life*.^23^ Excellent reviews of EMA for overall health behavior can be found in Persky, et. al, 2022^24^ and Morris et. al, 2020.^25^ A review of the advantages of EMA use in the SB population is available in Hensel et. al, 2023,^2^ Szymanski et. al, 2024,^12^ or Hensel et. al, 2025.^26^

Accordingly, the primary objectives of our paper were to provide preliminary data on:

1. How often YASB avoid ADL when they either experience *actual* UI and/or FI or when they are worried about *possible* UI and/or FI (Objective 1a) and the most commonly avoided activities in these situations (Objective 1b);
2. Associations between daily ADL avoidance and daily affect or incontinence anxiety (Objective 2);
3. Association between beginning-of-study HRQoL and daily ADL avoidance, as well as between daily ADL avoidance and end-of-study HRQoL (Objective 3).

## Methods

### Data and Participants

Participants were a young adults 18-27 years (N=23; 26.1% of all study-participants [N=88]) in a larger 30-day study (R21DK121355) study examining the feasibility of using EMA to understand daily incontinence experiences among adults with SB. All participants completed end-of-day EMAs that assessed UI- and FI-specific characteristics, including any ADL avoidance, and if relevant, the specific activities avoided. Additional methodological details in Hensel et. al, 2023^2^ and Szymanski et. al, 2024.^12^ The larger project was approved by the Institutional Review Board of Indiana University (#1907916729), and achieved excellent retention (97.5%), EMA completion (95.8%), data accuracy (98.6%). Participants rated acceptability high (>90% enjoyed the study, found protocol easy, felt supported, would participate again).^2,12^ Participant data are available in Table 1.

**Table 1.**
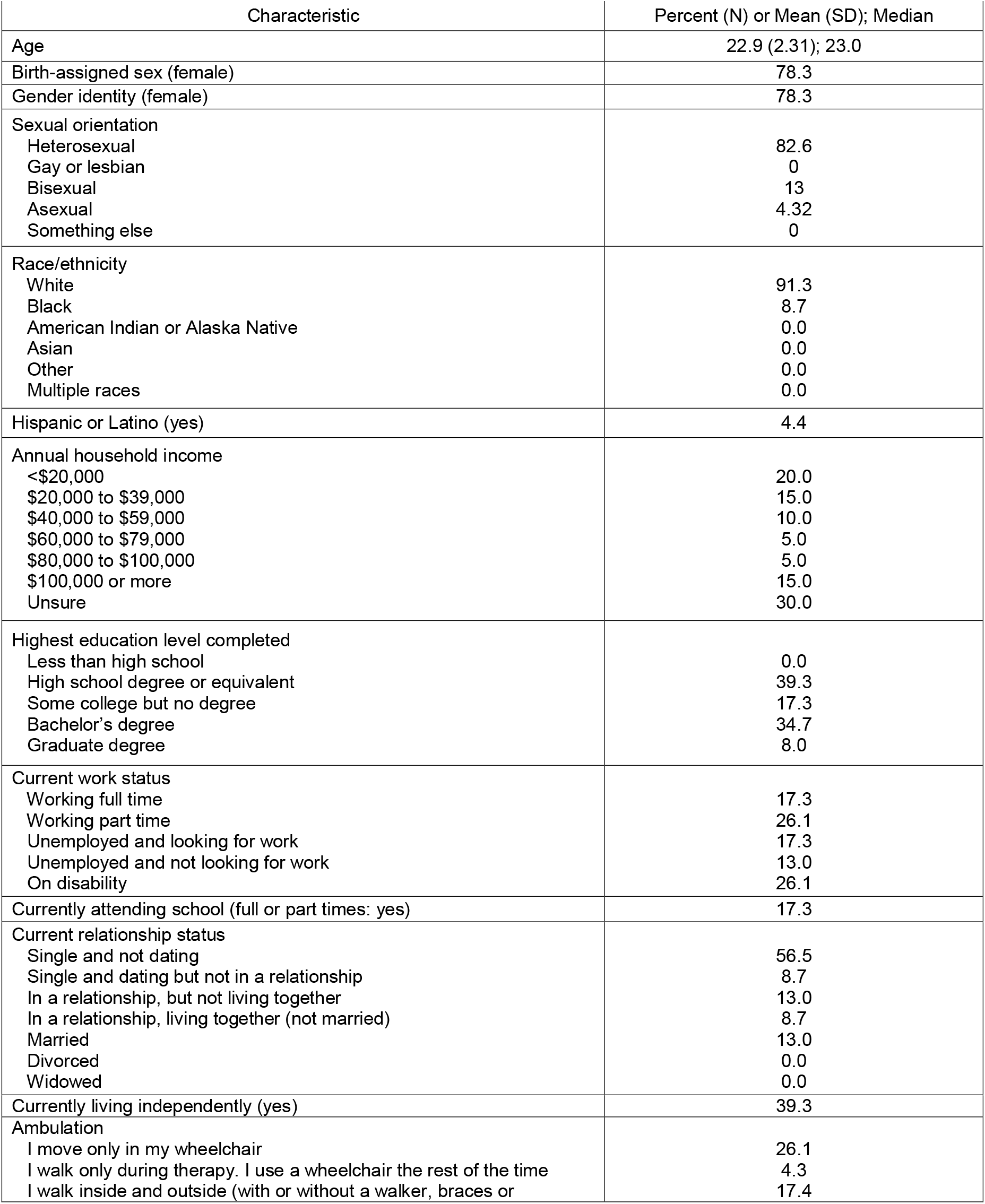
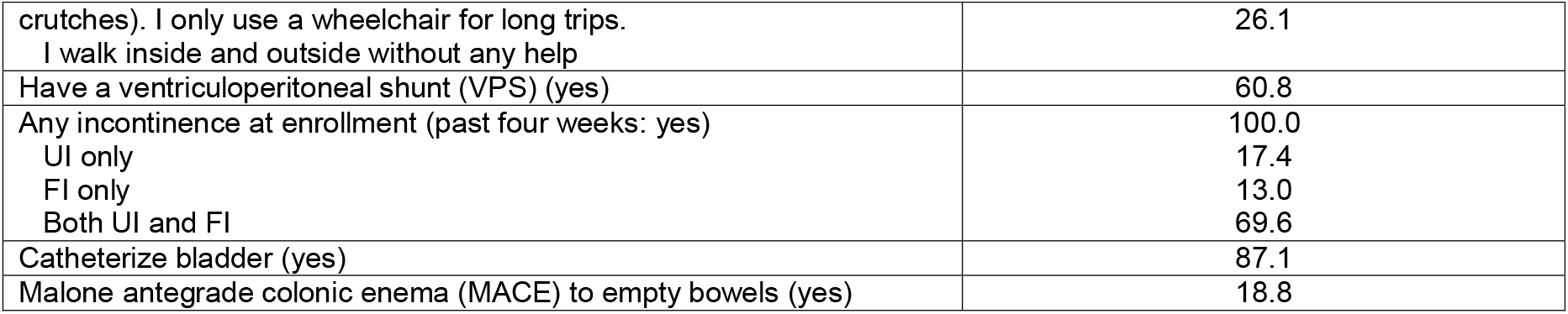
Demographic and background characteristics of young adults with Spina Bifida. Table 1. Characteristics of Young Adults with Spina Bifida (YASB).

### Measures

*ADL Avoidance*. Participants reported on any *activity avoidance with actual incontinence* (no/yes; separate item for UI and FI) and any *activity avoidance when worried about possible incontinence* (no/yes; separate item for UI and FI). An affirmative answer was followed by a question asking which of four basic (chores/housework, eating a meal/snack, drinking a beverage, going to a medical appointment) or ten instrumental (exercise, taking transportation, going to school/work, having sex, in person time with family/friends, doing a hobby, attending religious service, texted/called someone, watched TV/movies, took a nap/rested) activities they avoided (all: no/yes).

#### Daily Affect

We included positive mood (“happy” and “friendly”) and negative mood (“angry” and “irritable”) (PANAS; both additive indices of the two, 5-point Likert items [not at all to extremely]).^27^

#### Daily Incontinence Anxiety

We used a single 4-point Likert-type items that asked: “To what extent did you feel anxious about [incontinence type] today?” (not at all or very slightly, a little, moderately, extremely).

#### Health-related Quality of Life

In the enrollment and exit surveys of the study, participants completed the Health and Relationships (a person’s perception of health quality and connections with others) domain of the *Quality of Life Assessment in Spina Bifida for Adults*.^1^

### Statistics

We used descriptive statistics to characterize the daily frequency of ADL avoidance when YASB experience actual UI or FI or when they are worried about UI or FI (Objective 1a), as well as to identify the types of basic and instrumental ADL most commonly avoided (Objective 1b). Owing to the smaller number of EMAs available for analysis, comparisons between incontinence types were performed using a Wilcoxon Rank sum tests, which is a nonparametric equivalent for related samples.^28^ We used nonparametric Fisher’s Exact tests to examine median differences in positive mood, negative mood and incontinence anxiety on days when ADL were and were not avoided,^28^ and we used Spearman rank correlation coefficient to examine the preliminary associations between HRQoL and ADL avoidance (Objective 2).^29^ Finally, we used Spearman’s Rank Coefficient to assess the correlation between HRQoL at the beginning of the study and ADL avoidance, and between ADL avoidance and HRQoL at the end of the study (Objective 3).^30^

## Results

### Overall patterns

#### Overall daily urinary and fecal incontinence frequency

About six out of every ten days were associated with a report of any type of incontinence (57.5%; 370/643). The majority of these reports (63.7%: 236/370) were UI only. About one in ten (9.3%: 22/370) were FI only, and a third (30.2%: 112/370). Any UI was reported on 54.1% (348/643) of all study days. FI was noted on 20.8% (134/643) of all study days (*p*<.001).

*Objective 1a – Describe the frequency of daily ADL avoidance among YASB when they actually experienced UI or FI and ADL avoidance on non-incontinence days when they were worried about possible UI or FI*.

#### Urinary incontinence

YASB reported avoiding ADL with relatively the same frequency – about eight percent – on days when they were worried about possible UI (8.3%: 25/295) as they did on days when they actually experienced UI (8.9%: 31/348; *p*=0.546). This suggests slightly less than one out of every ten days with a real or perceived concern about UI involved the avoidance of at least one activity. As shown in Table 2, the median number of ADL avoided per day for both sources were one, ranging between no activities and two activities avoided around worry about UI and ranging between no activities and three activities avoided when UI occurred.

**Table 2.** Daily Reports of Activity of Daily Living Avoidance with Actual Incontinence or Worry About Possible Incontinence Among Young Adults with Spina Bifida.

|  | Worry Avoidance Type |  |  |  |  |  |  |  |  |  |  |  |
| --- | --- | --- | --- | --- | --- | --- | --- | --- | --- | --- | --- | --- |
|  | Worry about Urinary Incontinence | Actual Urinary Incontinence | Worry about Fecal Incontinence | Actual Fecal Incontinence |  |  |  |  |  |  |  |  |
| Total ALDs avoided per day (median) | 1 | 1 | 1 | 1 |  |  |  |  |  |  |  |  |
| Total ADLs avoided per day (range) | 0 - 2 | 0 - 3 | 0 - 2 | 0 - 4 |  |  |  |  |  |  |  |  |
| Basic ADL (%) |  |  |  |  |  |  |  |  |  |  |  |  |
| Chores/housework | 0.0 | 12.9 | 5.6 | 11.1 |  |  |  |  |  |  |  |  |
| Eating a meal | 44.0 | 16.1 | 55.6 | 55.6 |  |  |  |  |  |  |  |  |
| Drinking a beverage | 88.0 | 67.7 | 44.0 | 22.2 |  |  |  |  |  |  |  |  |
| Instrumental ADL (%) |  |  |  |  |  |  |  |  |  |  |  |  |
| Exercised | 44.0 | 19.3 | 11.1 | 18.5 |  |  |  |  |  |  |  |  |
| Attending a medical appointment | 0.0 | 0.0 | 0.0 | 3.7 |  |  |  |  |  |  |  |  |
| Took bus, train, car or plane | 12.0 | 3.2 | 0.0 | 3.7 |  |  |  |  |  |  |  |  |
| Went to school or work | 0.0 | 3.2 | 5.6 | 7.4 |  |  |  |  |  |  |  |  |
| Had sex | 0.0 | 19.6 | 11.1 | 14.8 |  |  |  |  |  |  |  |  |
| Spent time with family or friends | 4.0 | 12.9 | 27.8 | 33.3 |  |  |  |  |  |  |  |  |
| Did a hobby | 8.0 | 9.6 | 5.6 | 11.1 |  |  |  |  |  |  |  |  |
| Went to religious service | 0.0 | 3.2 | 0.0 | 3.7 |  |  |  |  |  |  |  |  |
| Texted, called someone or used the internet | 0.0 | 0.0 | 0.0 | 0.0 |  |  |  |  |  |  |  |  |
| Watched TV or movie | 0.0 | 0.0 | 0.0 | 3.7 |  |  |  |  |  |  |  |  |
| Took a nap or rested | 16.0 | 6.5 | 5.6 | 3.7 |  |  |  |  |  |  |  |  |
| Day-Level Characteristics | Median |  | Wilcoxon *p*-value | Median |  | Wilcoxon *p*-value | Median |  | Wilcoxon *p*-value | Median |  | Wilcoxon *p*-value |
|  | No | Yes |  | No | Yes |  | No | Yes |  | No | Yes |  |
| Positive mood (range: 2 - 10) | 6.0 | 5.0 | 0.119 | 6.0 | 6.0 | 0.234 | 6.0 | 5.0 | 0.003 | 6.0 | 6.0 | 0.051 |
| Negative mood (range: 2 - 10) | 3.0 | 4.0 | 0.101 | 3.0 | 4.0 | 0.001 | 3.0 | 3.0 | 0.054 | 3.0 | 4.0 | 0.001 |
| Incontinence anxiety (range: 1 - 5) | 1.0 | 3.0 | <.001 | 2.0 | 3.0 | <.001 | 1.0 | 3.0 | <.001 | 2.0 | 4.0 | <.001 |

ADL = activity of daily living; CI = Confidence Interval

#### Fecal incontinence

YASB noted ADL about five times less frequently on days when they were worried about FI (3.5%: 18/509) versus days when they actually experienced FI (20.2%: 27/134; *p*=0.002). These latter data suggest that one out of every five days with FI also involved activity avoidance. As shown in Table 2, the median number of ADL avoided per day for actual FI and worry about FI sources were one, ranging between no activities and two activities avoided around worry about FI and ranging between no activities and four activities avoided when FI occurred.

#### Supplementary analyses

We explored different comparisons *across* worry types. On 112 days when YASB reported both actual UI and FI, 12.5% (14/112) were associated with ADL avoidance for UI only and 7.1% (8/112) involved avoiding ADL around both UI and FI. No days (0.0%) were ADL avoidance for FI only (p=0.223 across the three groups). On 273 days when neither UI nor FI were reported, 1.8% (5/273) were concurrent with avoiding activities for worry about UI and 10.2% (25/273) for worry about FI only. No days had activity avoidance around worry for both UI and FI (p=0.567 across the three groups).

Daily ADL avoidance was significantly *more frequent* when YASB reported *actual* FI than when they reported *actual* UI (*p*<0.001). Conversely, daily ADL avoidance was significantly *more frequent* on days when YASB *worried* about UI as compared to when they *worried* about FI (*p*<0.001).

*Objective 1b – Describe the most commonly ADL YASB avoid they either experienced UI or FI or when they were worried about possible UI or FI.*

Table 2 displays the frequency of both basic and instrumental ADL avoidance, by actual incontinence and worry about incontinence. Of basic ADL, the most broadly endorsed activities avoided were eating and drinking. Of instrumental activities, the most commonly avoided activities were spending time with family or friends. Avoidance of eating a meal or snack was most frequent on days with actual UI (44.0%), with actual FI (55.6%) or worry about FI (55.6%). Drinking was most commonly avoided on days with worry about possible UI (88.0%) or with actual UI (67.7%), with worry about FI (44.0%) or with actual FI (22.2%). YASB reported avoiding exercises the most frequently when they were worried about UI (44.0%) (actual UI: 19.3% and actual FI: 18.5%). Finally, spending time with family and friends was most common on days with either worry about possible FI (27.8%) or with worry about possible FI (33.3%).

*Objective 2 – Understand preliminary median differences in positive mood, negative mood and incontinence anxiety on days with and without any ADL.*

As shown in Table 3, median UI anxiety was significantly *higher* on days when YASB reported ADL avoidance (vs. no avoidance) due to either worry about possible UI *or* experiencing actual UI (both *p*<.001). Median FI anxiety was also significantly higher on days when YASB reported ADL avoidance (vs. no avoidance) when they worried about possible FI *or* when they experienced actual FI (both *p*<.001). Negative mood was significantly *higher* on days when YASB avoided ADL both due to actual UI and actual FI (both *p*<.001) and positive mood was significantly *lower* on days when youth avoided ADL because they were worried about possible FI (*p*=.003).

**Table 3.** Daily Affect, Incontinence Anxiety and Activity of Daily Living Avoidance among Young Adults with Spina Bifida.

|  | Avoided any ADL – worried about UI |  |  | Avoided any ADL – actual UI |  |  |
| --- | --- | --- | --- | --- | --- | --- |
|  | Median |  | Fisher's<br>Exact<br>*p*-value | Median |  | Fisher's<br>Exact<br>*p*-value |
|  | No | Yes |  | No | Yes |  |
| Positive mood (range: 2 - 10) | 6.00 | 5.50 | 0.119 | 6.00 | 6.00 | 0.234 |
| Negative mood (range: 2 - 10) | 3.00 | 4.00 | 0.101 | 3.00 | 4.00 | 0.001 |
| Incontinence anxiety (range: 1 - 5) | 1.00 | 3.00 | <.001 | 2.00 | 3.00 | <.001 |
|  | Avoided any ADL – worried about FI |  |  | Avoided any ADL – actual FI |  |  |
|  | Median |  | Fisher's<br>Exact<br>*p*-value | Median |  | Fisher's<br>Exact<br>*p*-value |
|  | No | Yes |  | No | Yes |  |
| Positive mood (range: 2 - 10) | 6.00 | 5.00 | 0.003 | 6.00 | 6.0 | 0.051 |
| Negative mood (range: 2 - 10) | 3.00 | 3.00 | 0.054 | 3.00 | 4.00 | 0.001 |
| Incontinence anxiety (range: 1 - 5) | 1.00 | 3.00 | <.001 | 2.00 | 4.00 | <.001 |

ADL = Activity of Daily Living; UI = urinary incontinence; FI = fecal incontinence

*Objective 3 – Association of ADL with Health Related Quality of Life.*

All data for this objective are shown below; as used a Bonferroni to adjust *p*-values for multiple comparisons.

Higher HRQoL at the *start of the study* was significantly correlated with a lower number of days *during the study* on which YASB avoided ADL related to actual UI (Spearman’s Rank [SR] = −0.226; *p*<.001), actual FI (SR = −0.163; *p*=0.004) or worry about FI (SR = −0.190; *p*<.001). Starting HRQoL was not correlated with avoidance due to worry about UI. Put another way, higher HRQoL before beginning the study seemed to buffer – or reduce – the frequency of activity avoidance during the 30 days.

A higher number of days *during the study* on which YASB avoided ADL due to actual UI (SR = −0.309; *p*<.001) or actual FI (SR = −0.165; *p*=0.003) was significantly correlated with lower HRQoL at the *end of the study*. Similarly, YASB who reported a higher number of ADL avoidance days due to worry about UI (SR=-0.214; *p*<.001) or worry about FI (SR=-0.335; *p*<.001) *during the study* reported significantly *lower* HRQoL at the *end of the study*. Put another way, increased activity avoidance – for whatever motivation – over the 30 days reduced their HRQoL at the end of the month.

## Discussion

A central goal of caring for YASB is understanding any “clinically significant” impact incontinence may have on the function in their lives, including avoiding important daily activities. In the current paper, we used EMA to capture these avoidance events with greater precision to how YASB actually experienced them in “real life.” Ours is the first study to contribute preliminary understanding of how often ADL avoidance occurs in the context of worry about incontinence or actual incontinence, what activities YASB most commonly avoid, and explore the impact of mood, incontinence anxiety and HRQoL on activity avoidance. While previous studies broadly suggest that UI and FI create barriers to ADL participation,^3-5^ our data indicate that this avoidance on a daily basis may be even more heterogenous that previously thought. While additional data will be needed to optimize EMA as a clinical or research tool in this population, below we highlight preliminary findings that we think are particularly important for maximizing YASB well-being.

The first important idea is that both the overall occurrence of ADL avoidance and the specific activities YASB avoided varied depending on the both the incontinence source (e.g. UI vs. FI) and the avoidance source (e.g. worry about possible vs. actual incontinence). In other words, a dry day is not a day free of the negative effects of incontinence even though no incontinence occurs. While these statements may seem obvious, it is important to consider that most standardized clinical instruments screen for a single “usual” or “typical” actual incontinence outcome and do not capture worry about possible incontinence on days it does not occur.^31,32^ Such an approach is unlikely to capture the fluctuation we observed over a 30-day period and ignore the effects of worry about potential incontinence. For example, if we consider that the median number of ADL avoided per day – one activity – is the same across incontinence source and avoidance source, we could falsely assume that avoidance consistently occurs at low levels. However, a closer look at the data would suggest important nuances, such as that YASB avoid vital basic ADL like eating and drinking frequently, or that YASB often forgo exercise with worry about UI, but not with actual UI, or that YASB avoid social time with family and friends about six times as often when they have actual FI as compared to when they have actual UI. These pieces of information are likely important inputs for more personalized continence care.^12^ While the ideal EMA cadence and collection time to support clinical decision making remains to be established, this is an area worthy of further investigation. The opportunity to increase life participation – if even on a single activity – is important for all YASB, regardless of function level.^10,11^

We observed that daily subjective factors were associated with activity participation. Specifically, median daily negative mood and daily incontinence anxiety were significantly higher when YASB reported that they avoided activities in every incontinence situation: when they were worried about possible UI, when they were worried about FI, when they reported UI and when they reported FI. While studies suggest that depression and anxiety are common in people with SB,^33^ the role of affect in the improvement and exacerbation of incontinence symptoms remains unknown. The idea of targeting daily emotions alongside treating incontinence itself – perhaps in real time as part of just-in-time interventions – is an intriguing patient-centered approach yet unused in clinical settings. It may also be possible to include strengthening emotional management as part of self-management strategies.^34^ Clinicians may consider screening for mood to open discussions about ADL avoidance.^35^

Finally, our data suggested a preliminary association of cumulative ADL during the study with both beginning-*and* end-of-study reports of HRQoL. Specifically, *higher* HRQoL at the beginning of the study was significantly associated with *lower* ADL avoidance during the study, and *fewer* days of ADL during the study were associated with significantly *higher* HRQoL at the end of the study. On the one hand, these findings could suggest that building HRQoL serves as an important buffering effect to ongoing ADL avoidance in daily life. On the other hand, it could also be that lower activity avoidance reflects better condition management, increasing HRQoL. Disentangling the mechanism of these effects and their iterative clinical potential will be the focus of our future research, particularly in terms of how HRQoL could be leveraged to manage both short- and long-term well-being in those with SB.

There are methodological limitations associated with our study. First, we captured only 30 days of data from each YASB, which is likely too short of a time period to observe the “typical cycle” of ADL avoidance. This limited data volume also limited our ability to explore how other day-level factors (for example, objective incontinence markers like frequency or volume) may have been associated with ADL avoidance. While future work is planned to establish the optimal study length, a longer period – perhaps over six to twelve months – may better reflect “real life” YASB avoidance patterns. Additional daily observations would also permit a more extensive analysis of how contextual factors – like frequency or amount of incontinence, or past avoidance habits – impact daily avoidance. Future studies will focus on assessing both what an optimal data capture frame might be, and whether this frame should occur in a specific context. Second, while we did assess avoidance across nineteen different activities, there may be other ADL (e.g. basic hygiene or getting dressed) impacted by incontinence that we did not include. Relatedly, if an ADL were infrequent in a YASB’s life, a lack of avoidance endorsement could have indicated lack of general participation rather than avoidance due to incontinence. While no list of activities will likely represent every single YASB, future work should consider using YASB input as part of study planning to ensure a robust set of choices. Third, ongoing research should consider assessment of day-level factors we were unable to include – such as pain^36^ – that could have impacted activity participation. This and other ecological influences are planned as part of future studies in this area.

There are also sample limitations associated with the current data. First, our sample was primarily White, heterosexual, well-educated, and exhibited a high degree of living and mobility independence. Self-selection bias associated with these characteristics – particularly independence – could mean that potential participants more caregiver-dependent, or potential participants with greater-than-mild developmental delay would be less apt to submit the surveys. However, the participants in the larger study had reasonably similar gender and neurological characteristics to other samples from SB-focused research. Second, the larger feasibility study by design excluded individuals under 18 years. Inclusion of teenage participants are planned as part of future work, particularly given that the developmental roots of adult incontinence management – of which ADL avoidance is a part – are likely linked to experiences in adolescence. Third, all participants had a recent history of incontinence. We did not capture incontinence location (e.g. per stoma or urethra/anus), which could have impacted YASB activity avoidance.

## Conclusion

This study is the first to capture a snapshot of what activity avoidance around incontinence – due to either worry about possible incontinence or actual incontinence – among YASB. Our data suggest that avoidance experiences are likely more varied than originally thought, and perhaps more importantly, have associations with both daily affect and incontinence anxiety as well as longer term HRQoL. Longer studies will be certainly needed to both capture a wider section of YASB life, and to establish the ideal data volume to enhance clinical decision making, Nevertheless, we argue that the user-centered and minimally invasive structure of EMA is well suited for gathering data from all people with SB about how they navigate daily life with incontinence. In addition to symptom reporting, EMA could do double duty as a platform to encourage patient-clinician communication and to facilitate the delivery of ongoing, personalized therapies.^37^

## Data Availability

Raw data files and codebooks are stored with the Open Science Framework (https://osf.io/gefqx/)

## Abbreviations

SB: spina bifida
EMA: ecological momentary assessment
UI: urinary incontinence
FI: fecal incontinence ADL = activity of daily living

